# FINDING STUNTING HOTSPOT AREAS IN SEVEN MAJOR ISLANDS USING SPATIAL ANALYSIS: FOR THE ACCELERATION OF STUNTING PREVENTION IN INDONESIA

**DOI:** 10.1101/2021.03.31.21254736

**Authors:** Tiopan Sipahutar, Tris Eryando, Meiwita Paulina Budhiharsana, Kemal N. Siregar, Muhammad Nur Aidi, Minarto, Diah Mulyawati Utari, Martya Rahmaniati, Harimat Hendarwan

## Abstract

**Objectives:** To find stunting hotspots district/ cities in Indonesia in seven major islands in Indonesia.

**Method:** This is an ecological study that using aggregate data. We used data from The Basic Health Research Report of Indonesia 2018 and The Poverty Data and Information Report from the Central Bureau of Statistics (BPS) 2018. We analyzed 514 districts/ cities in Indonesia that spread out in 7 major Islands with 34 provinces. We used The Euclidean distance method to determine the neighborhood. Moran’s test was occupied to identify autocorrelation while Moran’s Scatter Plot particularly in the high-high quadrant was used to identify stunting hotspot areas.

**Result:** It was found that there is autocorrelation among districts/ cities in four major islands namely Sumatera, Java, Sulawesi, and Bali Nusa Tenggara Timur (NTT) Nusa Tenggara Barat (NTB). We identified 135 districts/ cities as stunting hotspot areas that spread in 14 provinces in four islands.

**Conclusion:** There is autocorrelation among districts/ cities in Sumatera, Java, Sulawesi, and Bali NTT NTB which resulted in 135 districts/ cities identified as stunting hotspots in four major islands in Indonesia

**Policy implication:** Provide information to the government in prioritizing stunting prevention areas in Indonesia in term of the acceleration of stunting prevention.

**Summary:** Strengths and limitations of this study:

a. The hypothesis which states that the prevalence of stunting in one area is associated with the prevalence of stunting in the neighboring area is a new method that should be considered to set the policy.
b. The study results can be used by the government to set priority areas for stunting interventions in Indonesia because so far, the government has made priorities based on stunting prevalence and weighted by poverty. Thus, by setting the priority areas, the funds required will be less compare if it executed in districts of Indonesia simultaneously.
c. The weakness in this study is the geographical differences of Indonesia regions given the vastness of the Indonesian territory. Some of the districts are separated by oceans and the size of the area is sometimes extremely different. It becomes difficult in determining the neighborhood definition method. Some regions will have no neighbors under certain conditions. Therefore, further research can be carried out with different methods of neighborhood definition.

## Introduction

The world is still struggling with stunting in children under five, until 2018 as many as 150.8 million children under five worldwide are stunted ^1^. Stunting is a condition of growth failure in children under five due to chronic malnutrition, so the child is too short for his age. This is a result of poor nutrition in-utero and in early childhood ^1–3^. The prevalence of stunting has tended to decline, but the decline is still slow ^1^.

Indonesia is the fourth largest stunting prevalence in the world ^4^ as in 2018 there are 30.8% of stunting prevalence in children under five^5^. Nationally, the prevalence of stunting has indeed decreased by 6.4% since 2013, but the distribution pattern of the prevalence of stunting at the district / city level in Indonesia from 2015 to 2017 appears that stunting in some areas has increased over a period of three years.

Stunting is caused by many factors, including which are closely related (directly related) to family and home environment factors ^6–18^, insufficient food intake, namely low quality of food, low quality of feeding and quality of drinking water ^6,7,19–26^, breastfeeding ^27–30^ and infections ^17,19,25,26,31 32^. Stunting is also related to social and community factors such as the economic and political situation ^33–36^, health services, education, culture, food and agriculture systems ^37^ and related infrastructure of water, sanitation and environment ^38–42^.

Measurement of height by age at two years is the best predictor of human capital while stunting is associated with low human capital ^43^. Poor fetal growth or stunting at two years of age can cause permanent damage in terms of shortness in adulthood, low achievement in school, reducing income at adulthood and reducing birth weight of children. Children who are undernourished at up to two years of age and gain weight rapidly in childhood and in adulthood are at high risk for chronic nutrition-related illness ^43–49^. The effects of malnutrition will span at least three generations as shown by the significant relationship between grandmother height and birth weight of children born to women in the five cohorts ^43^.

At the end of 2017, the National Strategy for the Acceleration of Stunting Prevention 2018-2024 was issued ^50^. Stunting reduction in Indonesia is indeed a big challenge considering that Indonesia is a large country, consisting of 17,504 islands with 34 provinces and 514 district / city. Indonesia is the largest country in Southeast Asia, which has regional characteristics, socio-cultural characteristics, behavior, poverty levels that vary from island to island and even between districts / cities in a province. These factors should be taken into account in making stunting directives / policies / programs throughout Indonesia.

Spatial analysis in stunting context is still not widely used in Indonesia and may not even be used as a decision support system in making policies or programs, both at the national and regional levels. Therefore, Indonesia needs information on nutrition data which is analyzed by considering the regional context given the vastness of Indonesia’s territory and its different regional variations. The high prevalence of stunting and large gaps in many areas, and limited funds require the central and regional governments to prioritize the types of intervention and intervention areas as well as the need for quick action in order to meet the national target (19% of children under five by 2024) ^51^ dan Global World Health Assembly (40% pada tahun 2025) ^52^.

This study aims to find stunting hotspots area in Indonesia to provide information to the national, provincial and district governments in prioritizing stunting prevention areas in Indonesia in each of the major islands in Indonesia, namely Sumatra Island; Java; Kalimantan; Sulawesi; Nusa Tenggara Timur, Nusa Tenggara Barat, Bali; Maluku; and Papua using data from Basic Health Research Report 2018 as the main source of information. Thus, the government can decide the priority area of stunting intervention due to the acceleration of stunting prevention in Indonesia.

## Methods

This is an ecological study using aggregate data. Basic Health Research Report of Indonesia 2018 is the main data source in this research. Basic Health Research is nationally representative survey that provides stunting status in under five children in all districts/ cities in Indonesia. The units of analysis in this study are districts / cities throughout Indonesia as much as 514 districts / cities. There are 514 districts/ cities all in Indonesia that spread out in 7 major Islands namely Sumatera (10 provinces, 154 districts/ cities), Java (6 provinces, 119 districts/ cities), Kalimantan (5 provinces, 56 districts/ cities), Sulawesi (6 provinces, 81 districts/ cities), Bali Nusa Tenggara Timur Nusa (NTT) Tenggara Barat (NTB) (3 provinces, 41 districts/ cities), Maluku (2 provinces, 21 districts/ cities), and Papua (2 provinces, 42 districts/ cities).

The Euclidean Distance Method was used to determine spatial weighted. The neighborhood definition is if the distance among area within a radius of 1 degree or equivalent to 111 Kilometer (km) in accordance with Euclidean definition. We used Moran’s Test to prove whether there is autocorrelation among districts / cities in each island with a significance level of 0,05. The null hypothesis for autocorrelation is *I*=0. The hotspot areas were determined by Moran’s Scatter Plot. The areas that clustered in high-high quadrant will be referred as stunting hotspot areas which means that high stunting prevalence area surrounded by high stunting prevalence areas as well. The rationale of this determination is since there is autocorrelation among neighboring areas based on stunting prevalence so it is not enough to intervene in one area with a high prevalence of stunting, instead all clustered area should be intervened. Missing data is not allowed in this research; it was filled by calculating the mean of neighboring areas data. We did Anderson Darling (AD) test, Durbin-Watson (DW), VIF multicollinearity, and Breusch Pagan (BP) test prior to Moran Test to test respectively the residual normality, independency, multicollinearity, and homoscedasticity assumptions with α= 5%. There were no patients or public involvement in this research. All data used in this research are taken from public domain. We used software R i386 3.6.1 to run the analysis and Tableau Public 2020 to create the map.

## Result

The result of normality, independency, homoscedasticity, and multicollinearity assumption of stunting prevalence residual is fulfilled as shown in Table 1. There is significant autocorrelation **among** districts/ cities based on stunting prevalence in Sumatera, Java, Sulawesi, and Bali NTT NTB while on the other hand, all districts/ cities in Kalimantan, Maluku and Papua have no autocorrelation with their neighboring areas.

**Table 1.** Statistic Test Result and Moran’s Index Value of Each Island.

| Islands | Statistic test result/ p-value | Moran's Index value/ p-value |
| --- | --- | --- |
| Sumatera | AD: 0.537 9 (p-value: 0.166)<br>DW: 2.052 (p-value: 0.832)<br>BP: 3.562 (p-value: 0.829)<br>VIF all variables < 10 | I: 0.299 (p-value: 1.522e-10) |
| Java | AD: 0.714 (p-value: 0.0609)<br>DW: 1.754 (p-value: 0.154)<br>BP: 10.253 (p-value: 0.419)<br>VIF all variables < 10 | I: 0.105 (p-value: 1.246e-06) |
| Kalimantan | AD: 0.42 (p-value: 0.315)<br>DW: 1.868 (p-value: 0.56)<br>BP: 4.851 (p-value: 0.773)<br>VIF all variables < 10 | I: 0.104 (p-value: 0.073) |
| Sulawesi | AD: 0.4696 (p-value: 0.241)<br>DW: 1.868 (p-value: 0.48)<br>BP: 6.604 (p-value: 0.678)<br>VIF all variables < 10 | I: 0.303 (p-value: 2.038e-09) |
| Bali NTT NTB | AD: 0.669 (p-value: 0.075)<br>DW: 1.727 (p-value: 0.34)<br>BP: 11.809 (p-value: 0.298)<br>VIF all variables < 10 | I: 0.633 (p-value: 4.127e-15) |
| Maluku | AD: 0.149 (p-value: 0.956)<br>DW: 2.393 (p-value: 0.516)<br>BP: 7.608 (p-value: 0.574)<br>VIF all variables < 10 | I: -0.128 (p-value: 0.4103) |
| Papua | AD: 0.243 (p-value: 0.751)<br>DW: 2.655 (p-value: 0.02)<br>BP: 5.851 (p-value: 0.664)<br>VIF all variables < 10 | I: 0.126 (p-value: 0.55) |

**Table 2.**
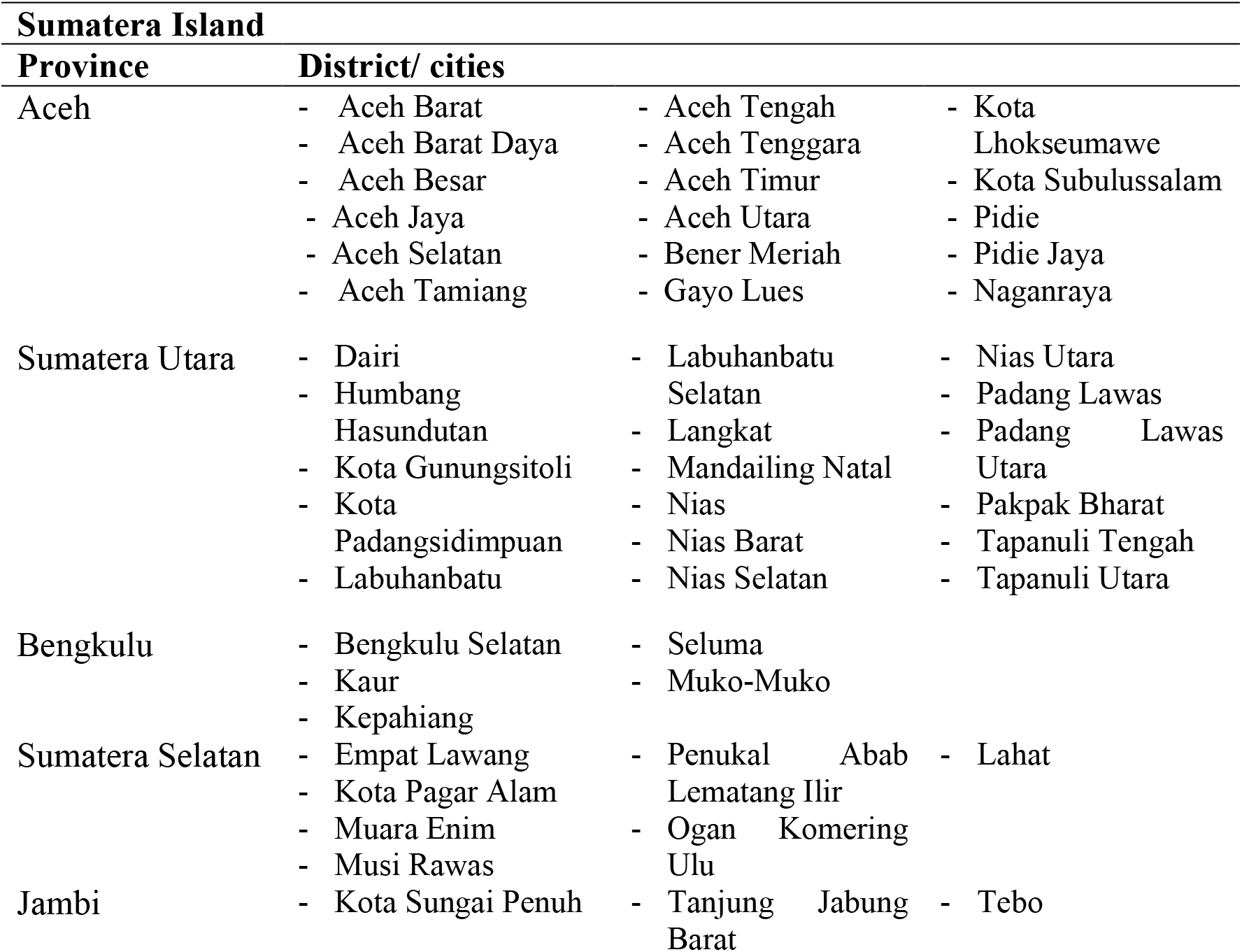
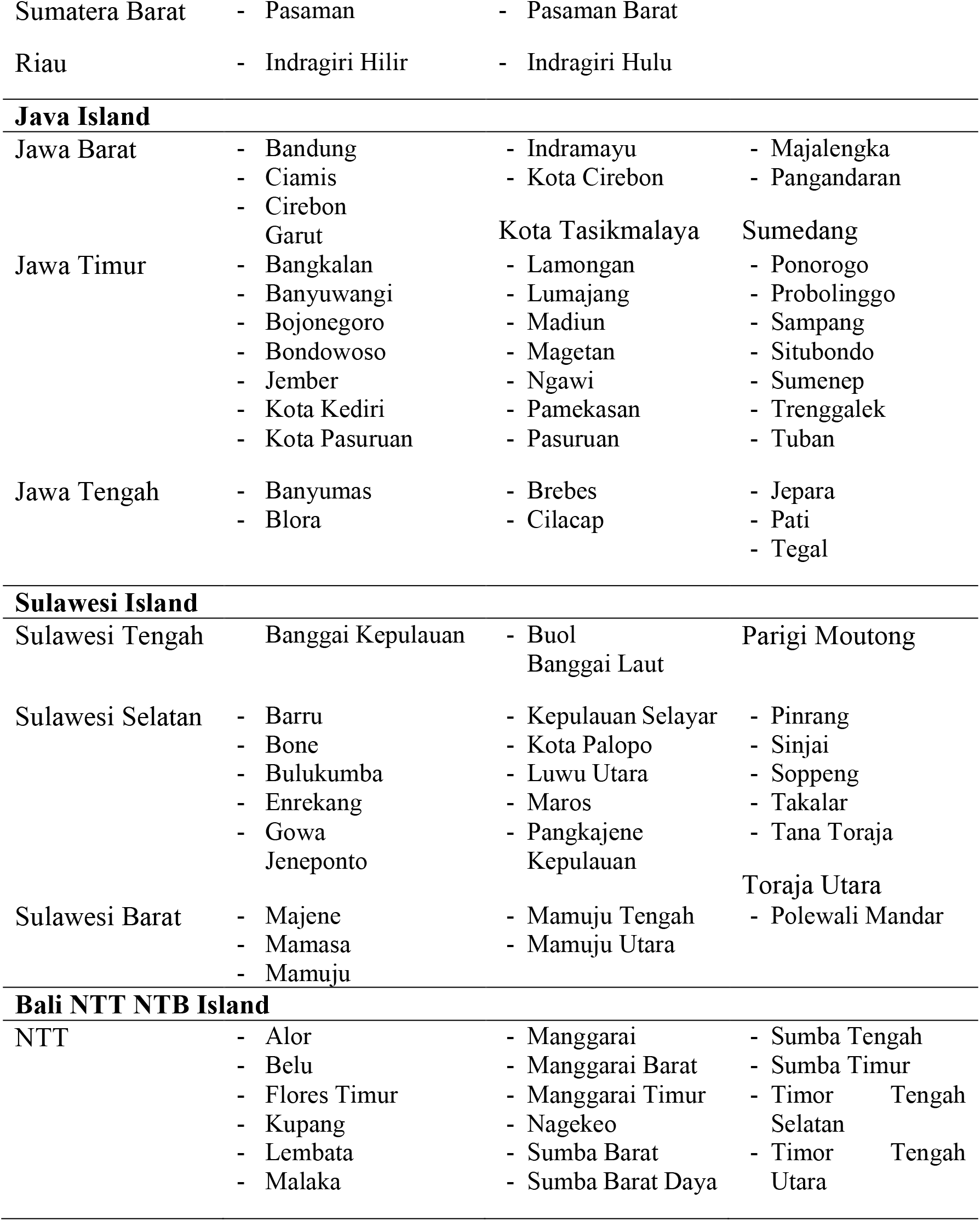
The Stunting Hotspot District/ Cities in Indonesia in 2018.

The Moran’s Scatter Plot of Sumatera, Java, Sulawesi, and Bali NTT NTB are shown in Picture 1.

**Picture 1.**
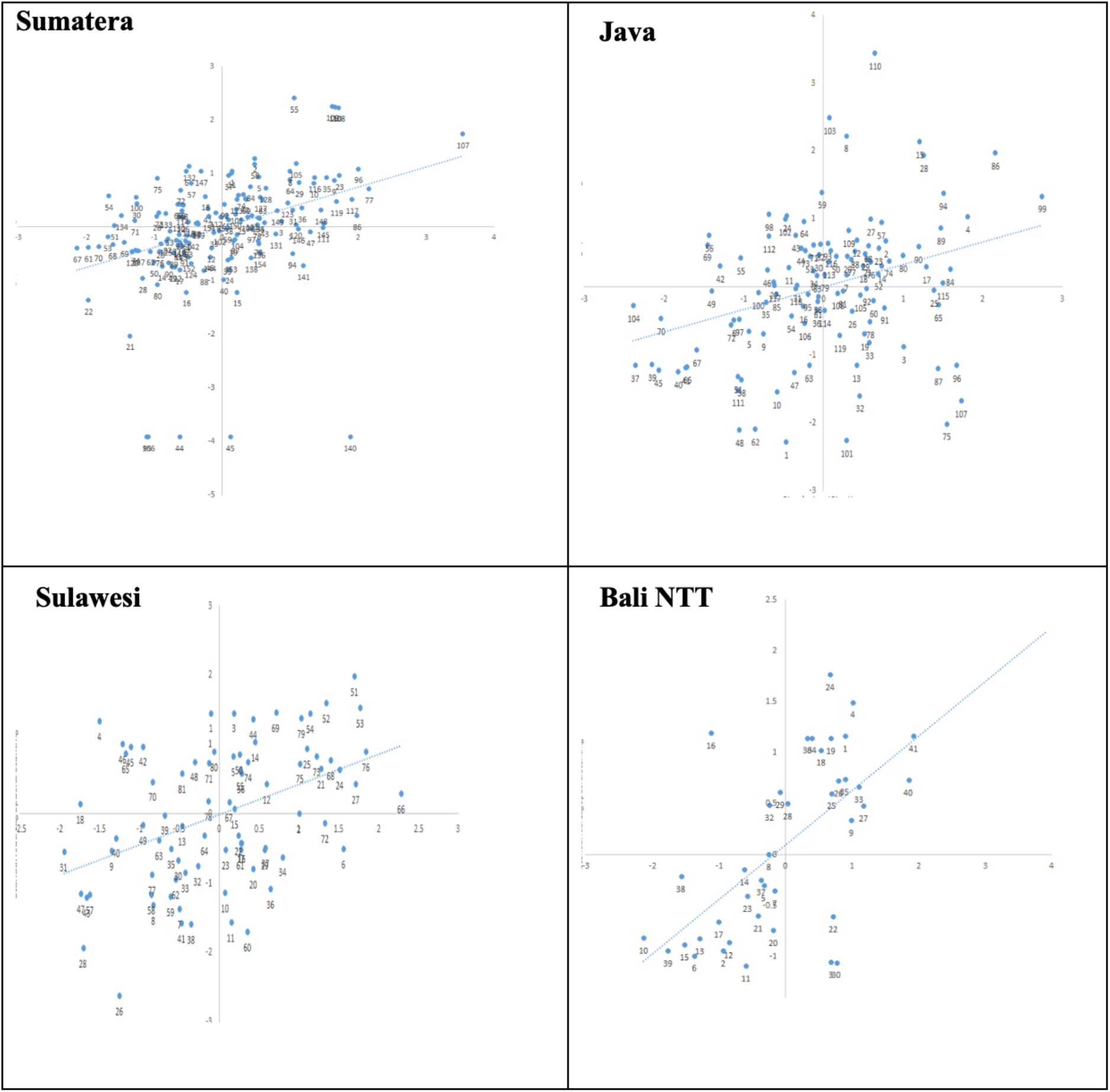
Moran’s Scatter Plot of Sumatera, Java, Sulawesi and Bali NTT NTB Islands.

The hotspot areas are all districts/ cities that clustered in high-high quadrat in Moran’s Scatter Plot (Picture 1.). We identified that there are 135 hotspot districts/ cities that spread in 14 provinces in four islands. Picture 2. describes the geographic distribution of hotspot areas in Indonesia.

**Picture 2.**
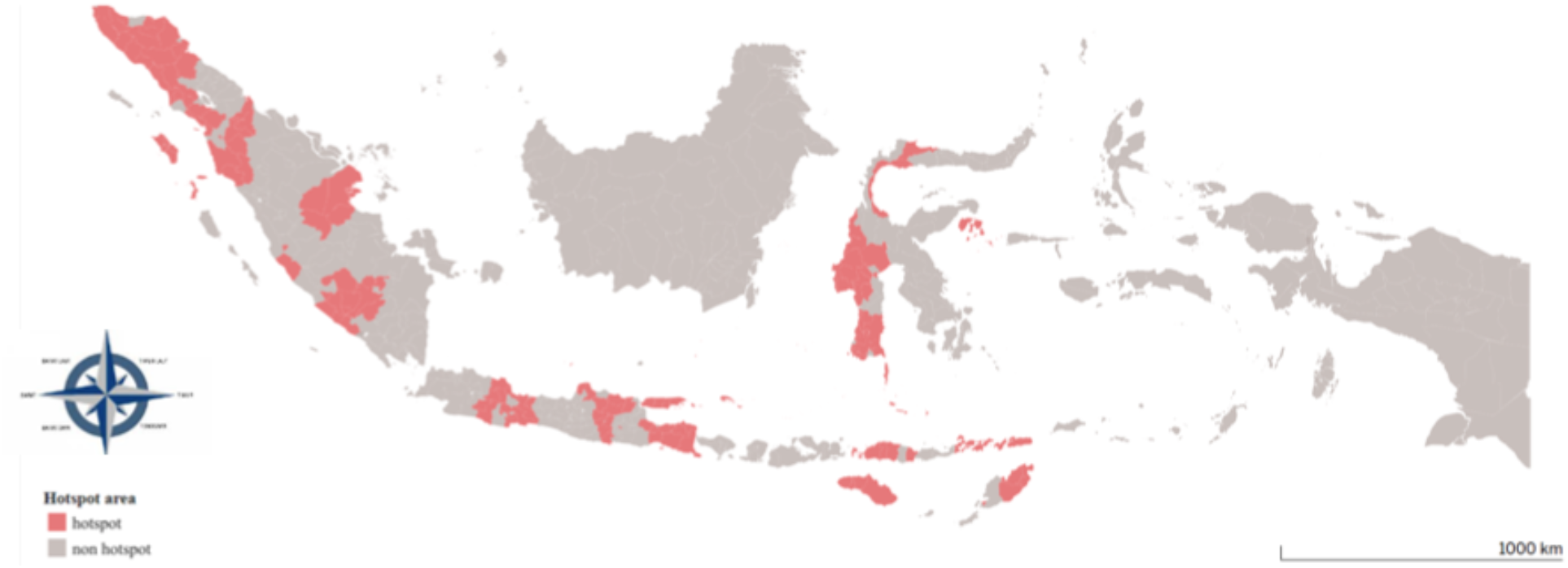
Stunting Hotspot Area of Indonesia Based on Spatial Analysis 2018.

## Discussion

The autocorrelation in Sumatera, Java, Sulawesi and Bali NTT NTB Island indicates that the prevalence of stunting -whether high or low -in one district / city does not occur randomly however it is related to the prevalence of stunting in the surrounding districts / cities. The purpose of spatial autocorrelation is indeed to prove whether the stunting prevalence among districts are related to the stunting prevalence levels of the neighboring districts/ cities (not randomly occurred) ^53^. The attribute value of a variable from an area tends to be the same or almost the same as a region close to it compared to a region further away. This is based on the basic concept of geography (Tobler I’s Law) which states “everything is related to everything else, but near things are more related than distant thing"^53–57^. The autocorrelation that has been proven to occur in these four islands can make the pattern of stunting interventions more tailored based on specific regions. Some previous study in other country showed the similar result such as in India, Africa, and Peru^585942^

Spatial autocorrelation among districts/ cities could not be identified in Kalimantan, Maluku and Papua. Kalimantan Island tends to be similar to Papua Island in terms of area size; two of the five provinces in Kalimantan Island are the largest provinces in Indonesia. Thus, the distance between one district to another district becomes further away due to the size of one district. Papua Island is the largest island in Indonesia where the population lives in groups according to their ethnicity and the population is not evenly distributed^60^. Maluku has a geographic situation that is slightly different from Papua and Kalimantan where the districts in Maluku are separated by the ocean. From the explanation above, it is known that the size of a district and the geographical shape that is separated by the ocean greatly affects the distance between one area and its neighboring area. This then affects the presence or absence of spatial autocorrelation based on the prevalence of stunting. The theory says that the attribute value of a variable in a region tends to be the same or almost the same as a region closer to it than a region farther away^53–57^. In spatial analysis context, autocorrelation is the similarity varying with the distance between locations and how this variation is affected by the distance between locations^61^.

The determination of priority areas for stunting interventions in Indonesia is currently only based on the high prevalence of stunting and weighted by the percentage of poverty in the region so that then this determination method only to arrange the intervention time instead of making priorities. However, using the spatial analysis method, it will be determined the priority areas in Indonesia based on identified hotspot in a certain period of time. The rationale for this is that the prevalence of stunting in districts / cities is related to the prevalence of stunting in nearby areas so that the interventions should be aimed at areas that constitute one cluster.

Some limitation might occur in this study such as ecological study that is prone to ecological fallacy where aggregate data representing areas are applied at the individual level^5362^. We did estimation to fill the missing data. The weakness of data estimation is that it cannot completely / cannot be ascertained to represent the real situation.

## Conclusion

Indonesia is currently planning to accelerate stunting prevention to reduce the prevalence of stunting to 14% by 2024. Considering the vast area of Indonesia with all the differences in regional characteristics, it is necessary to have priority areas for intervention. Spatial analysis can help to determine priority areas by looking at hotspots generated from Moran’s Scatter Plot, especially areas that are in the high-high quadrant. This study found that there are 135 districts / cities of stunting hotspots in Indonesia out of 514 districts / cities throughout Indonesia. Hotspots are spread across four major islands, namely Sumatra, Java, Sulawesi, and Bali NTT NTB. All of these hotspots were recommended to the government to become priority areas for stunting intervention. By knowing that there is autocorrelation between district/ city and that stunting does not occur randomly in the four regions, then intervention programs should be carried out in these hotspot clusters.

Stunting hotspots are not found in Kalimantan, Maluku, and Papua because there is no autocorrelation in those three islands. Furthermore, it may be necessary to carry out a similar study using different spatial weighted.

## Data Availability

All data that have been used in this study are public domain. People can access all data freely without any requirements.

https://www.litbang.kemkes.go.id/laporan-riset-kesehatan-dasar-riskesdas/

https://www.bps.go.id/pressrelease/2019/07/15/1629/persentase-penduduk-miskin-maret-2019-sebesar-9-41-persen.html

## Data Sharing Statement

Data are available online from the Central Bureau of Statistics (BPS) and Health Ministry of Indonesia. Data inserted in BPS report and Province Basic Health Research Report.

## Contributorship Statement

All authors have made substantial contribution to this research and have approved the final manuscript. TS contributed on all steps of research concept, design, writing, and data interpretation. TE, MPB, KNS, MNA, Mi, DMU, MR, HH were contributed on concept, interpretation, and review.

## Ethics Approval and Consent to Participate

The study was based on data available in public domain; therefore, no ethical issue is involved.

## References

1. UNICEF/WHO/World Bank Group. Levels and Trends in Child Malnutrition-Key Findings of the 2018edition.; 2018. doi:10.1016/S0266-6138(96)90067-4

2. ACC/SCN. Fourth Report on the World Nutrition Situation. Geneva ACC/SCN Collab with IFPRI. Published online 2000:140.

3. Onis de M, Branca F. Childhood stunting: A global perspective. Matern Child Nutr. 2016;12. doi:10.1111/mcn.12231

4. Tim Nasional Percepatan Penanggulangan Kemiskinan (TNP2K). Gerakan Nasional Pencegahan Stunting dan Kerjasama Kemitraan Multi Sektor. Published online 2017:1-42.

5. Badan Penelitian Dan Pengembangan Kesehatan Kementerian Kesehatan RI. Riset Kesehatan Dasar 2018.; 2019.

6. Beal T, Tumilowicz A, Sutrisna A, Izwardy D, Neufeld LM. A review of child stunting determinants in Indonesia. Matern Child Nutr. 2018;14(4). doi:10.1111/mcn.12617

7. Torlesse H, Cronin AA, Sebayang SK, Nandy R. Determinants of stunting in Indonesian children: Evidence from a cross-sectional survey indicate a prominent role for the water, sanitation and hygiene sector in stunting reduction. BMC Public Health. 2016;16(1). doi:10.1186/s12889-016-3339-8

8. Sinha B, Taneja S, Chowdhury R, et al. Low-birthweight infants born to short-stature mothers are at additional risk of stunting and poor growth velocity: Evidence from secondary data analyses. Matern Child Nutr. 2018;14(1):1–9. doi:10.1111/mcn.12504

9. K.G.D, R.J. C. Does birth spacing affect maternal or child nutritional status? A systematic literature review. Matern Child Nutr. 2007;3(3):151–173.

10. Rahman MS, Howlader T, Masud MS, Rahman ML. Association of low-birth weight with malnutrition in children under five years in Bangladesh: Do mother’s education, socio-economic status, and birth interval matter? PLoS One. 2016;11(6). doi:10.1371/journal.pone.0157814

11. Saxton J, Rath S, Nair N, et al. Handwashing, sanitation and family planning practices are the strongest underlying determinants of child stunting in rural indigenous communities of Jharkhand and Odisha, Eastern India: a cross-sectional study. Matern Child Nutr. 2016;12(4). doi:10.1111/mcn.12323

12. Rah JH, Cronin AA, Badgaiyan B, Aguayo V, Coates S, Ahmed S. Household sanitation and personal hygiene practices are associated with child stunting in rural India: A cross-sectional analysis of surveys. BMJ Open. 2015;5(2). doi:10.1136/bmjopen-2014-005180

13. Mostafa I, Naila NN, Mahfuz M, Roy M, Faruque ASG, Ahmed T. Children living in the slums of Bangladesh face risks from unsafe food and water and stunted growth is common. Acta Paediatr Int J Paediatr. 2018;107(7):1230–1239. doi:10.1111/apa.14281

14. Spears D, Ghosh A, Cumming O. Open Defecation and Childhood Stunting in India: An Ecological Analysis of New Data from 112 Districts. PLoS One. 2013;8(9):1–10. doi:10.1371/journal.pone.0073784

15. Shamah-Levy T, Nasu LC, Moreno-Macias H, Monterrubio-Flores E, Avila-Arcos MA. Maternal Characteristics Determine Stunting in Children of Less than Five Years of Age Results from a National Probabilistic Survey. Clin Med Pediatr. 2008;1:pCMPed.S1019. doi:10.4137/cmped.s1019

16. Semba RD. Effect of parental education on child stunting – Author’s reply. Lancet. 2008;371(9627):1837. doi:10.1016/s0140-6736(08)60793-x

17. Ikeda N, Irie Y, Shibuya K. Determinants of reduced child stunting in Cambodia: analysis of pooled data from three Demographic and Health Surveys. Bull World Health Organ. 2013;91(5):341–349. doi:10.2471/blt.12.113381

18. Danaei G, Andrews KG, Sudfeld CR, et al. Risk Factors for Childhood Stunting in 137 Developing Countries: A Comparative Risk Assessment Analysis at Global, Regional, and Country Levels. PLoS Med. 2016;13(11). doi:10.1371/journal.pmed.1002164

19. Kusumawati E, Rahardjo S, Sari HP. Model Pengendalian Faktor Risiko Stunting pada Anak Usia di Bawah Tiga Tahun. J Kesehat Masy. 2013;9(3):249–256.

20. Panjwani A, Heidkamp R. Complementary Feeding Interventions Have a Small but Significant Impact on Linear and Ponderal Growth of Children in Low-and Middle-Income Countries: A Systematic Review and Meta-Analysis. J Nutr. Published online 2017. doi:10.3945/jn.116.243857

21. Aguayo VM, Menon P. Stop stunting: Improving child feeding, women’s nutrition and household sanitation in South Asia. Matern Child Nutr. 2016;12. doi:10.1111/mcn.12283

22. Simanjuntak YB, Haya M, Suryani D, Ahmad CA. Early Inititation of Breastfeeding and Vitamin A Supplementation with Nutritional Status of Children Aged 6-59 Months. Natl Public Heal J. 2018;12(3):107–113. doi:10.21109/kesmas.

23. M’Kaibi FK, Steyn NP, Ochola SA, Du Plessis L. The relationship between agricultural biodiversity, dietary diversity, household food security, and stunting of children in rural Kenya. Food Sci Nutr. 2017;5(2). doi:10.1002/fsn3.387

24. Smith LC, Haddad L. Reducing Child Undernutrition: Past Drivers and Priorities for the Post-MDG Era. World Dev. 2015;68(1). doi:10.1016/j.worlddev.2014.11.014

25. Asfaw M, Wondaferash M, Taha M, Dube L. Prevalence of undernutrition and associated factors among children aged between six to fifty nine months in Bule Hora district, South Ethiopia. BMC Public Health. 2015;15(1). doi:10.1186/s12889-015-1370-9

26. Kinyoki DK, Berkley JA, Moloney GM, Kandala NB, Noor AM. Predictors of the risk of malnutrition among children under the age of 5 years in Somalia. Public Health Nutr. 2015;18(17):3125–3133. doi:10.1017/S1368980015001913

27. Cetthakrikul N, Topothai C, Suphanchaimat R, Tisayaticom K, Limwattananon S, Tangcharoensathien V. Childhood stunting in Thailand: When prolonged breastfeeding interacts with household poverty. BMC Pediatr. 2018;18(1):1–10. doi:10.1186/s12887-018-1375-5

28. Jiang Y, Su X, Wang C, et al. Prevalence and risk factors for stunting and severe stunting among children under three years old in mid-western rural areas of China. Child Care Health Dev. 2015;41(1). doi:10.1111/cch.12148

29. Badriyah L, Syafiq A. The Association Between Sanitation, Hygiene, and Stunting in Children Under Two-Years (An Analysis of Indonesia’s Basic Health Research, 2013). Makara J Heal Res. 2017;21(2):35–41. doi:10.7454/msk.v21i2.6002

30. Rachmi CN, Agho KE, Li M, Baur LA. Stunting coexisting with overweight in 2·0-4·9-year-old Indonesian children: Prevalence, trends and associated risk factors from repeated cross-sectional surveys. Public Health Nutr. 2016;19(15). doi:10.1017/S1368980016000926

31. Akombi BJ, Agho KE, Hall JJ, Merom D, Astell-Burt T, Renzaho AMN. Stunting and severe stunting among children under-5 years in Nigeria: A multilevel analysis. BMC Pediatr. 2017;17(1). doi:10.1186/s12887-016-0770-z

32. WHO. Childhood Stunting?: Context, Causes and Consequences WHO Conceptual framework. 2013;9(September):27–45.

33. Vollmer S, Harttgen K, Subramanyam MA, Finlay J, Klasen S, Subramanian S V. Association between economic growth and early childhood undernutrition: Evidence from 121 Demographic and Health Surveys from 36 low-income and middle-income countries. Lancet Glob Heal. 2014;2(4):e225–e234. doi:10.1016/S2214-109X(14)70025-7

34. Pongou R, Ezzati M, Salomon JA. Household and community socioeconomic and environmental determinants of child nutritional status in Cameroon. BMC Public Health. 2006;6:1–19. doi:10.1186/1471-2458-6-98

35. Jonah CMP, Sambu WC, May JD. A comparative analysis of socioeconomic inequities in stunting: A case of three middle-income African countries. Arch Public Heal. 2018;76(1):1–15. doi:10.1186/s13690-018-0320-2

36. Fenske N, Burns J, Hothorn T, Rehfuess EA. Understanding child stunting in India: A comprehensive analysis of socio-economic, nutritional and environmental determinants using additive quantile regression. PLoS One. 2013;8(11). doi:10.1371/journal.pone.0078692

37. Hagos S, Hailemariam D, WoldeHanna T, Lindtjørn B. Spatial heterogeneity and risk factors for stunting among children under age five in Ethiopia: A Bayesian geo-statistical model. PLoS One. 2017;12(2):1–18. doi:10.1371/journal.pone.0170785

38. Hagos S, Lunde T, Mariam DH, Woldehanna T, Lindtjørn B. Climate change, crop production and child under nutrition in Ethiopia; A longitudinal panel study. BMC Public Health. 2014;14(1):1–9. doi:10.1186/1471-2458-14-884

39. Chikhungu LC, Madise NJ. Seasonal variation of child under nutrition in Malawi: Is seasonal food availability an important factor? Findings from a national level cross-sectional study. BMC Public Health. 2014;14(1):1–11. doi:10.1186/1471-2458-14-1146

40. Kandala N-B, Madungu TP, Emina JB, Nzita KP, Cappuccio FP. Malnutrition among children under the age of five in the Democratic Republic of Congo (DRC): does geographic location matter? BMC Public Health. 2011;11(1):261. doi:10.1186/1471-2458-11-261

41. Kinyoki DK, Berkley JA, Moloney GM, Odundo EO, Kandala NB, Noor AM. Environmental predictors of stunting among children under-five in Somalia: Cross-sectional studies from 2007 to 2010. BMC Public Health. 2016;16(1):1–10. doi:10.1186/s12889-016-3320-6

42. Hernández-Vásquez A, Tapia-López E. [Chronic Malnutrition among Children under Five in Peru: A Spatial Analysis of Nutritional Data, 2010-2016]. Rev Esp Salud Publica. 2017;91(May). http://www.ncbi.nlm.nih.gov/pubmed/28509895

43. Victora CG, Adair L, Fall C, et al. Maternal and child undernutrition: consequences for adult health and human capital. Lancet. 2008;371(9609):340–357. doi:10.1016/S0140-6736(07)61692-4

44. Dewey KG, Begum K. Long-term consequences of stunting in early life. Matern Child Nutr. 2011;7(SUPPL. 3):5–18. doi:10.1111/j.1740-8709.2011.00349.x

45. E.D.L. R, G.V.A.D. F, C.A. V, et al. Associations of stunting in early childhood with cardiometabolic risk factors in adulthood. PLoS One. 2018;13(4):1–13. doi:10.1371/journal.pone.0192196 LK -http://findit.library.jhu.edu/resolve?sid=EMBASE&issn=19326203&id=doi:10.1371%2Fjournal.pone.0192196&atitle=Associations+of+stunting+in+early+childhood+with+cardiometabolic+risk+factors+in+adulthood&stitle=PLoS+ONE&title=PLoS+ONE&volume=13&issue=4&spage=&epage=&aulast=Rolfe&aufirst=Emanuella+De+Lucia&auinit=E.D.L.&aufull=Rolfe+E.D.L.&coden=POLNC&isbn=&pages=-&date=2018&auinit1=E&auinitm=D.L.

46. Hoddinott JJ.R. B, J.A. M, et al. Adult consequences of growth failure in early childhood. Am J Clin Nutr. Published online 2013:1170-1178. doi:http://dx.doi.org/10.3945/ajcn.113.064584

47. Semba RD, Shardell M, Sakr Ashour FA, et al. Child Stunting is Associated with Low Circulating Essential Amino Acids. EBioMedicine. 2016;6. doi:10.1016/j.ebiom.2016.02.030

48. Grantham-McGregor S, Cheung YB, Cueto S, Glewwe P, Richter L, Strupp B. Developmental potential in the first 5 years for children in developing countries. Lancet. 2007;369(9555):60–70. doi:10.1016/S0140-6736(07)60032-4

49. Casale D, Desmond C, Richter L. The association between stunting and psychosocial development among preschool children: A study using the South African Birth to Twenty cohort data. Child Care Health Dev. 2014;40(6):900–910. doi:10.1111/cch.12143

50. Tim Nasional Percepatan Penanggulangan Kemiskinan Republik Indonesia (TNP2K). Strategi Nasional Percepatan Pencegahan Stunting 2018-2024. 2018;(November):1–32.

51. Bappenas. Rancangan Teknokratik Rencana Pembangunan Jangka Menengah Nasional 2020 - 2024?: Indonesia Berpenghasilan Menengah - Tinggi Yang Sejahtera, Adil, Dan Berkesinambungan.; 2019. doi:10.1017/CBO9781107415324.004

52. WHO. Global Nutrition Targets 2025 Stunting Policy Brief.; 2014. doi:10.1057/9781137477699_6

53. Souris M. Epidemiology and Geography. John Wiley & Sons, Ltd; 2019.

54. Fischer MM, Wang J. Spatial Data Analysis Models, Methods and Techniques. Springer Briefs in Regional Science; 2011. doi:10.1007/978-3-642-21720-3

55. Grekousis G. Spatial Analysis Methods and Practice?: Describe, Explore, Explain through GIS.; 2020.

56. Bhunia, G S; Shit PK. Geospatial Analysis of Public Health. Springer Nature Switzerland; 2019.

57. Waller, Lance A; Gotway CA. Applied Spatial Statistics for Public Health Data. Jhon Wiley & Sons, Inc; 2004. doi:10.1198/jasa.2005.s15

58. Khan J, Mohanty SK. Spatial heterogeneity and correlates of child malnutrition in districts of India. BMC Public Health. 2018;18(1):1–13. doi:10.1186/s12889-018-5873-z

59. Alemu ZA, Ahmed AA, Yalew AW, Birhanu BS. Non random distribution of child undernutrition in Ethiopia: Spatial analysis from the 2011 Ethiopia demographic and health survey. Int J Equity Health. 2016;15(1):1–11. doi:10.1186/s12939-016-0480-z

60. Bappenas. Analisis Wilayah Dengan Kemiskinan Tinggi.; 2018.

61. Fotheringham S, Rogerson P. The SAGE Handbook of Spatial Analysis. Sage Publication Ltd; 2009. doi:10.4135/9780857020130

62. Gerstman BB. Epidemiology Kept Simple. Third Edit. John Wiley & Sons, Ltd; 2013.

